## Supplementary material for "Temporo-spatial cellular atlas of the regenerating alveolar niche in idiopathic pulmonary fibrosis": Suppl Fig n Tables legend MedRxv

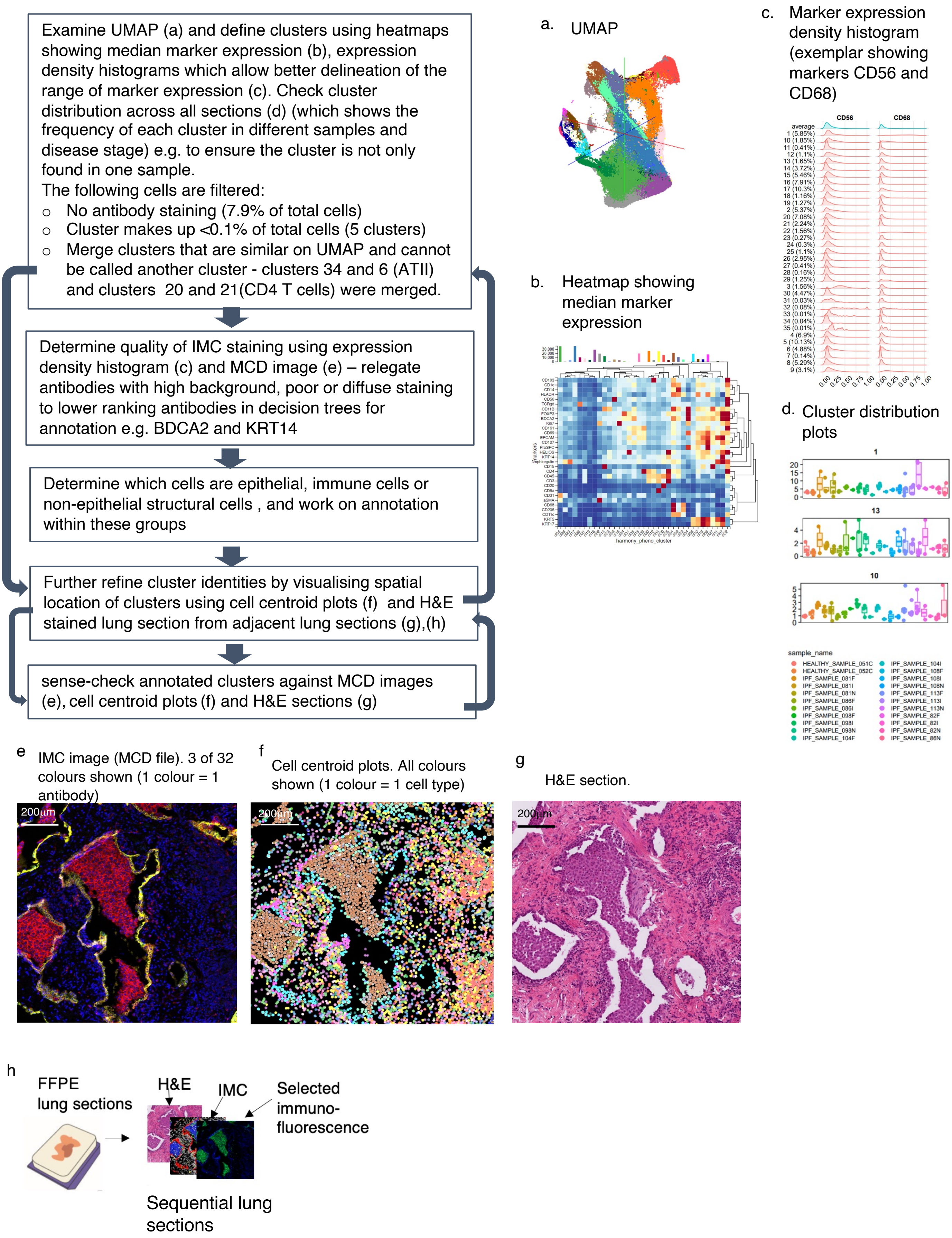

**Suppl Figure 1**

i

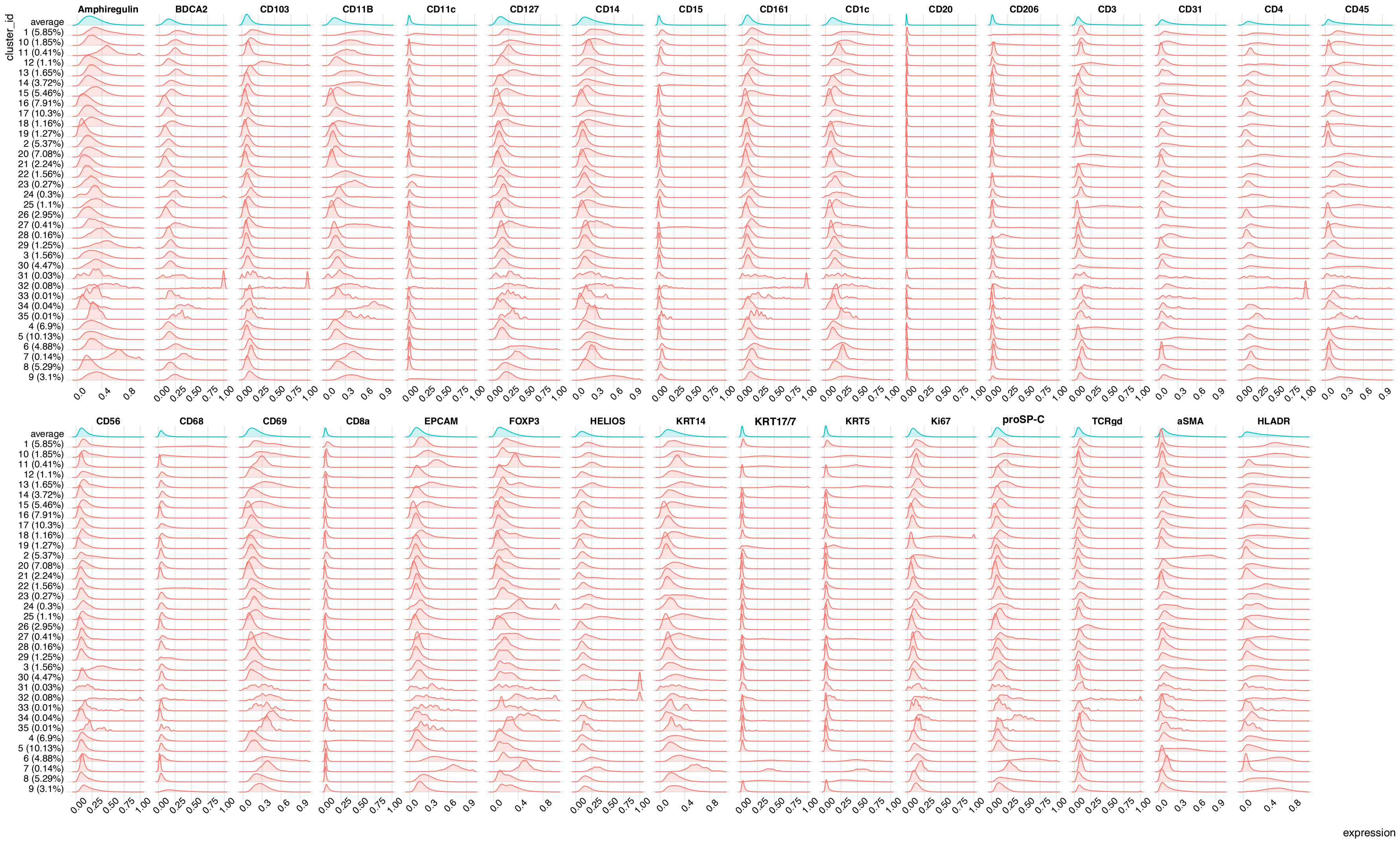

Cluster names found in Suppl Table 4

**Suppl Figure 1**

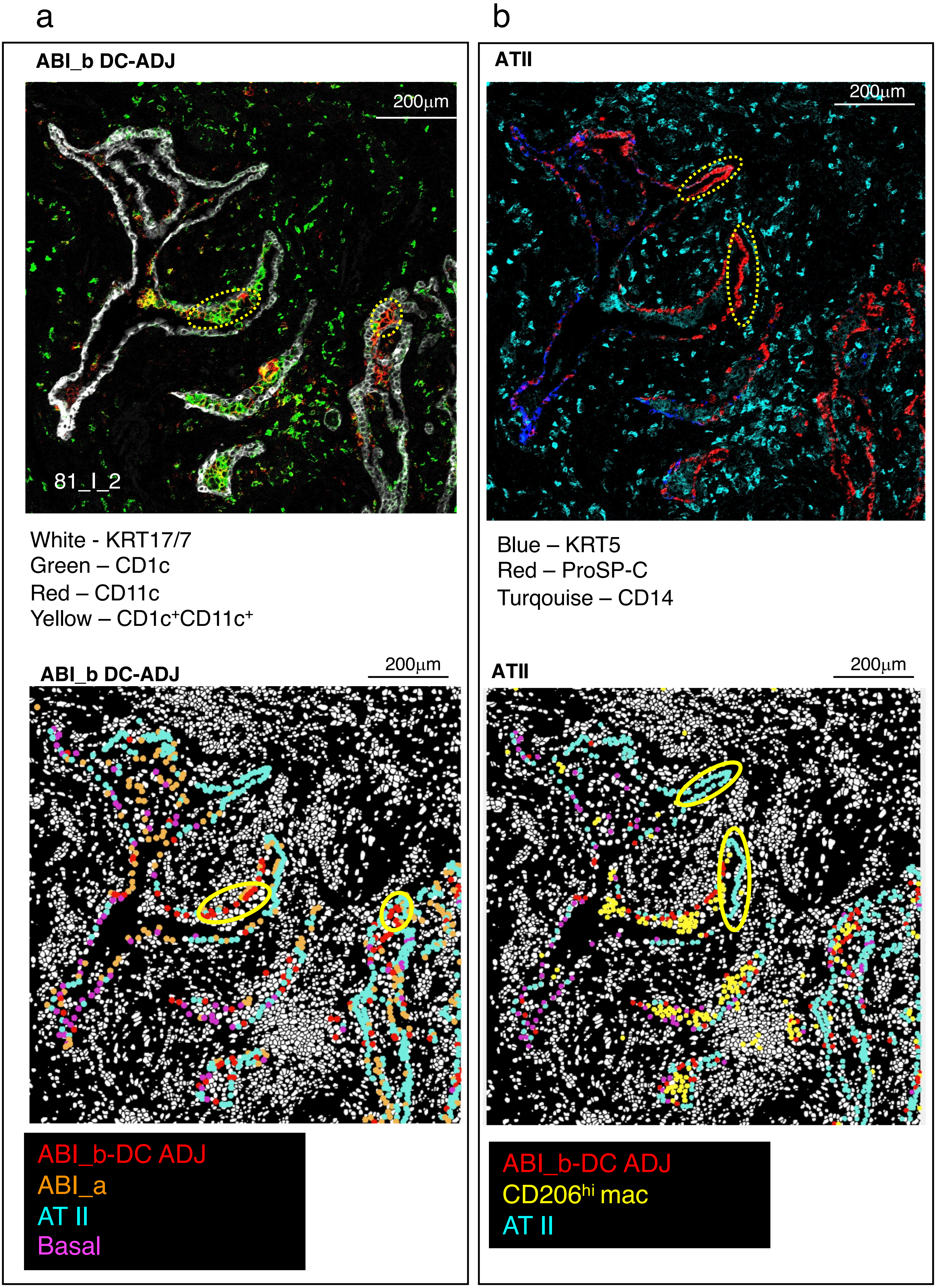

**Suppl Figure 2**

a. **Habermann AC et al Sci Adv 2020 (IPF lung tissue) (single cell transcriptomic study)**
Authors termed KRT5$^{-}$KRT17$^{lo}$ cells as alveolar intermediates. There is also a KRT5$^{neg}$ transitional AT II cell cluster.

**Adams TS et al Sci Adv 20202 (IPF lung tissue) (single cell transcriptomic study)**
Authors termed aberrant basaloid (AB) or KRT5$^{-}$ basaloid cells as alveolar intermediates.

**Kathiriya JJ et al Nat Cell Biol 2021(lung organoid studies) (gene expression)**
See Kathriya's Fig 7a. Authors called alveolar intermediates KRT5$^{neg}$ ABIs although there may be very low expression of KRT5 on these ABIs from their Fig 7a.

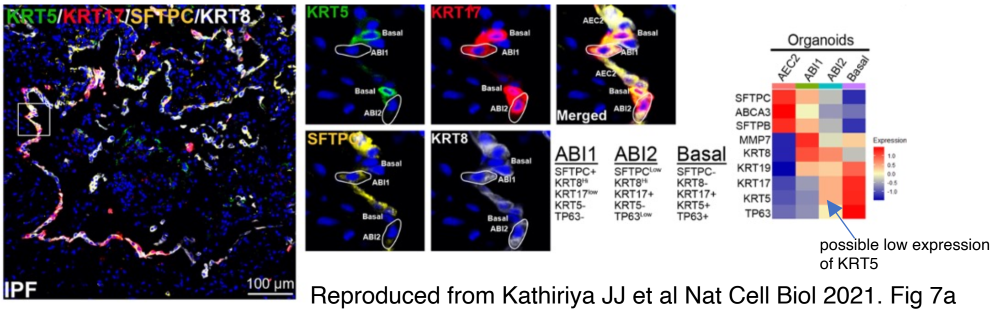

Reproduced from Kathiriya JJ et al Nat Cell Biol 2021. Fig 7a

**Weeratunga P (our paper) (IPF lung tissue)(IMC, protein)**
We termed alveolar intermediates:
ABI_a - KRT5$^{neg-lo}$proSP-C$^{neg-lo}$
ABI_b - KRT5$^{neg-lo}$ proSP-C$^{lo}$
ABI_b - DC ADJ - KRT5$^{neg-lo}$ proSP-C$^{lo}$

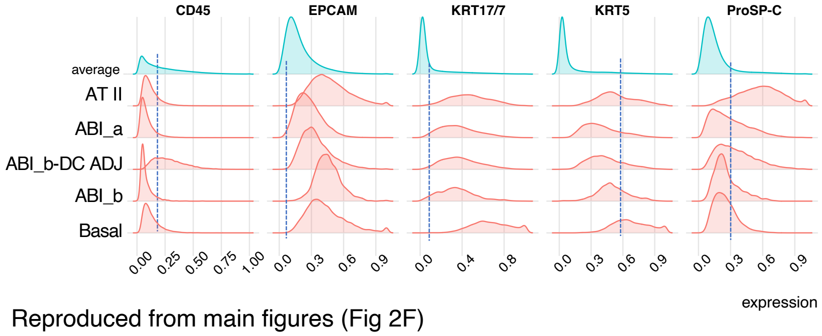

Reproduced from main figures (Fig 2F)

**Suppl Figure 3**

b. **Habermann AC et al Sci Adv 2020. Gene expression of KRT5, KRT17, KRT7 and SPFTC (proSP-C)**

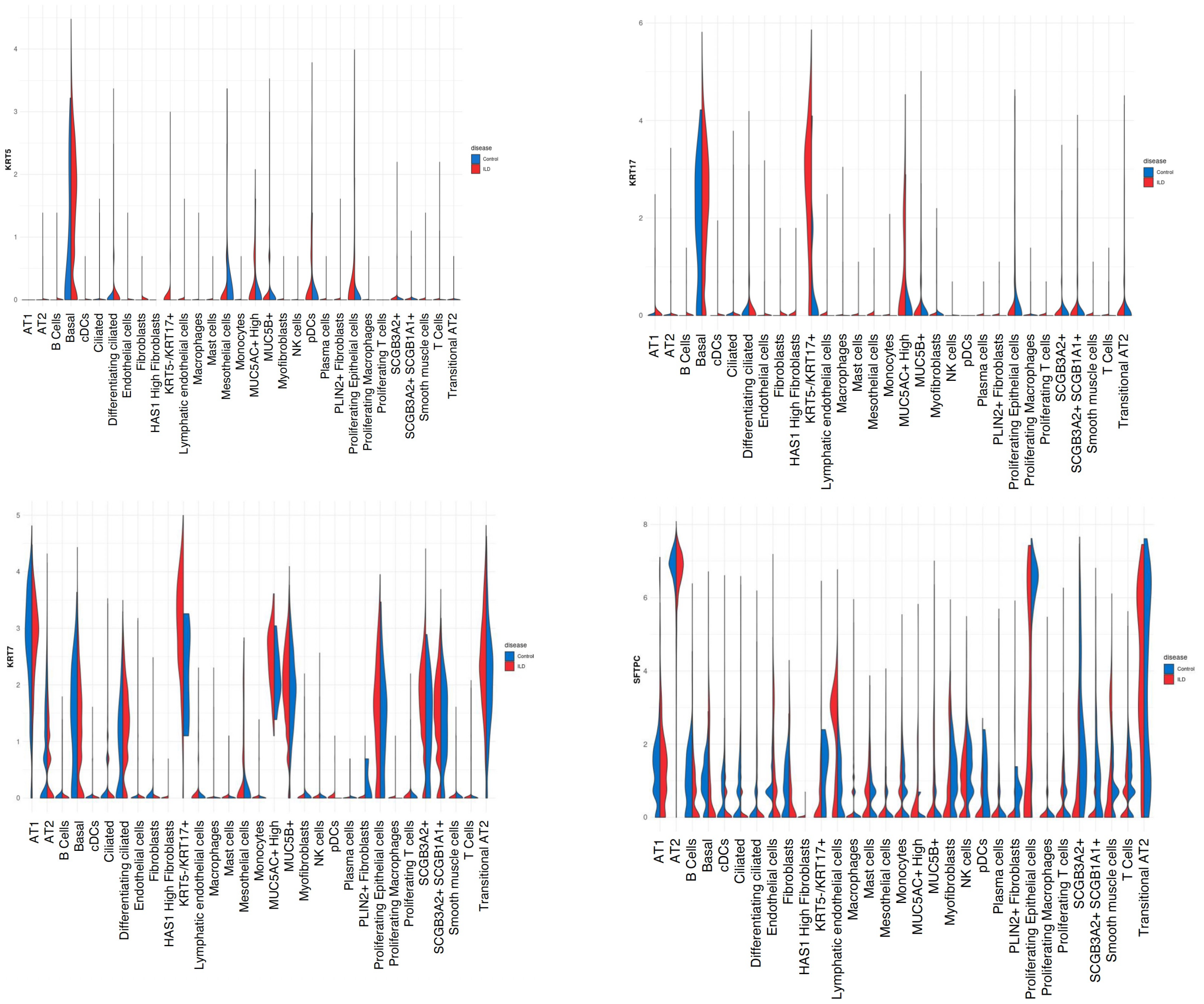

**Suppl Figure 3**

C. **Adams TS et al Sci Adv 20202**

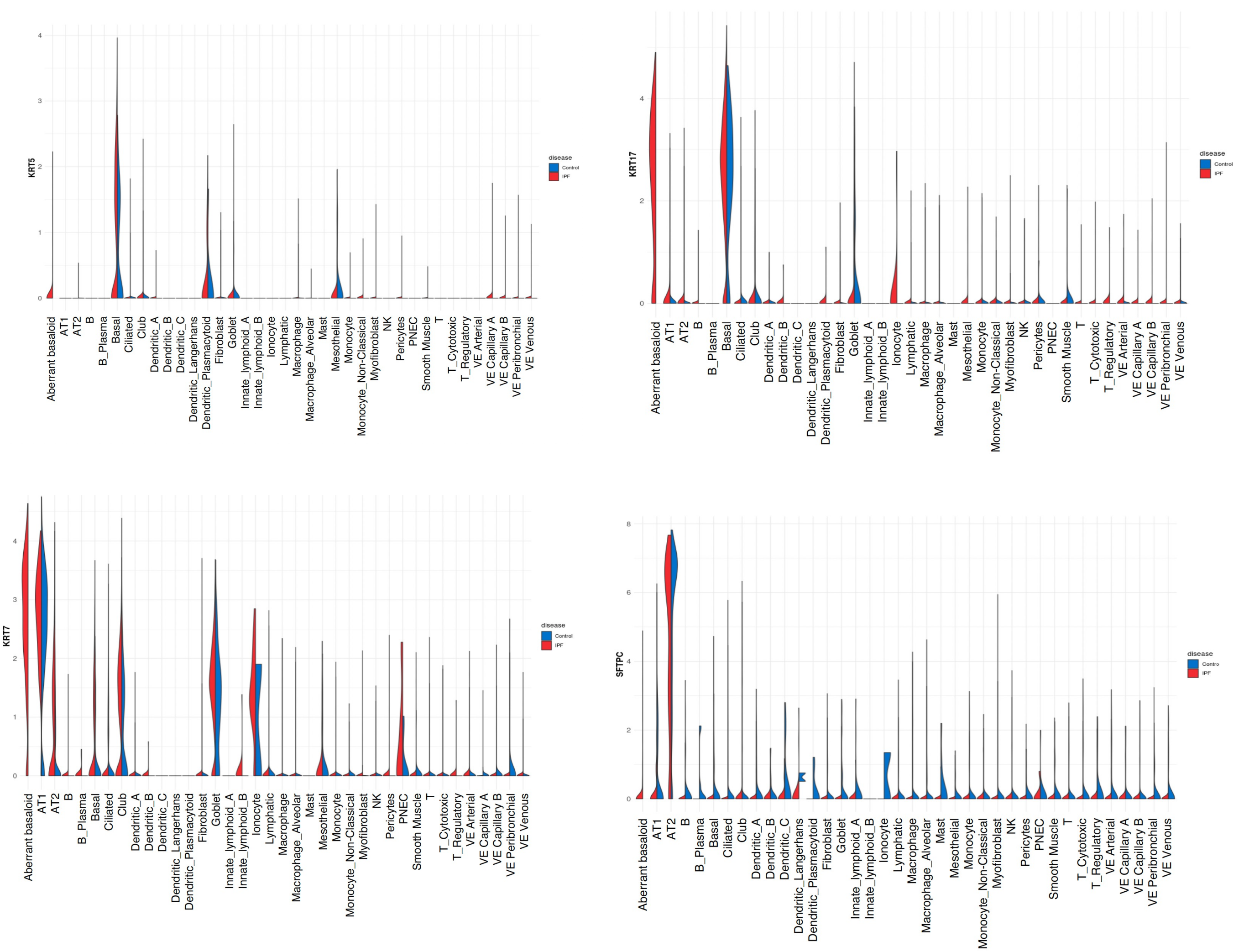

**Suppl Figure 3**

d. Combined data from Suppl Fig 3B and C shows that at gene level,

- ATI cells are $KRT5^{-}KRT17^{-}KRT7^{hi}$ $proSP\text{-}C^{lo}$
- ATII cells are $KRT5^{-}KRT17^{-}KRT7^{mid}$ $proSP\text{-}C^{hi}$
- KRT5-KRT17 AB cells are $KRT5^{-/lo}KRT17^{hi}KRT7^{hi}$ $proSP\text{-}C^{lo}$
- Transitional ATII cells are $KRT5^{-}KRT17^{lo}KRT7^{hi}$ $proSP\text{-}C^{mid\text{-}hi}$

*However, note that at single cell transcriptomic level, ATI and ATII derivation from lung digest is not a complete picture due to technical difficulties of retrieving these cells, in particular, ATI cells*

**Proposed terms of equivalence**
**for our protein-led alveolar-basaloid intermediates vs Habermann, Adams and Kathiriya**

**ABI_a =**

- Kathiriya's ABI 1 (possibly also ABI 2)
- Adam's AB
- Habermann's $KRT5^{-}KRT17^{lo}$ subset

**ABI_b =**

- Kathiriya's ABI 2 (possibly also ABI 1)
- Adam's AB
- Habermann's $KRT5^{-}KRT17^{lo}$ subset

e.

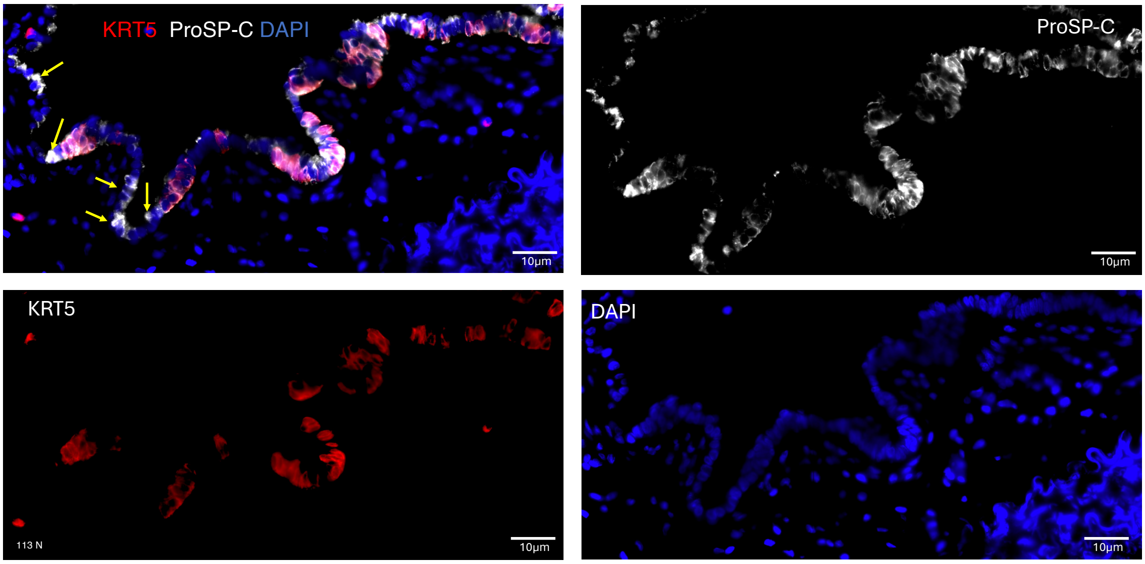

Yellow arrows – Single positive staining - $ProSp\text{-}C^{+}$ $KRT5^{neg}$ (AT II cells). All other cells double positive for KRT5 and ProSp-C (ABI_a and ABI_b)

***Weeratunga P (our paper) (IPF lung tissue)(IMC, protein)***
*We termed alveolar intermediates:*
*ABI_a - $KRT5^{neg\text{-}lo}proSP\text{-}C^{neg\text{-}lo}$*
*ABI_b - $KRT5^{neg\text{-}lo}$ $proSP\text{-}C^{lo}$*
*ABI_b - DC ADJ - $KRT5^{neg\text{-}lo}$ $proSP\text{-}C^{lo}$*

**Suppl Figure 3**

f.

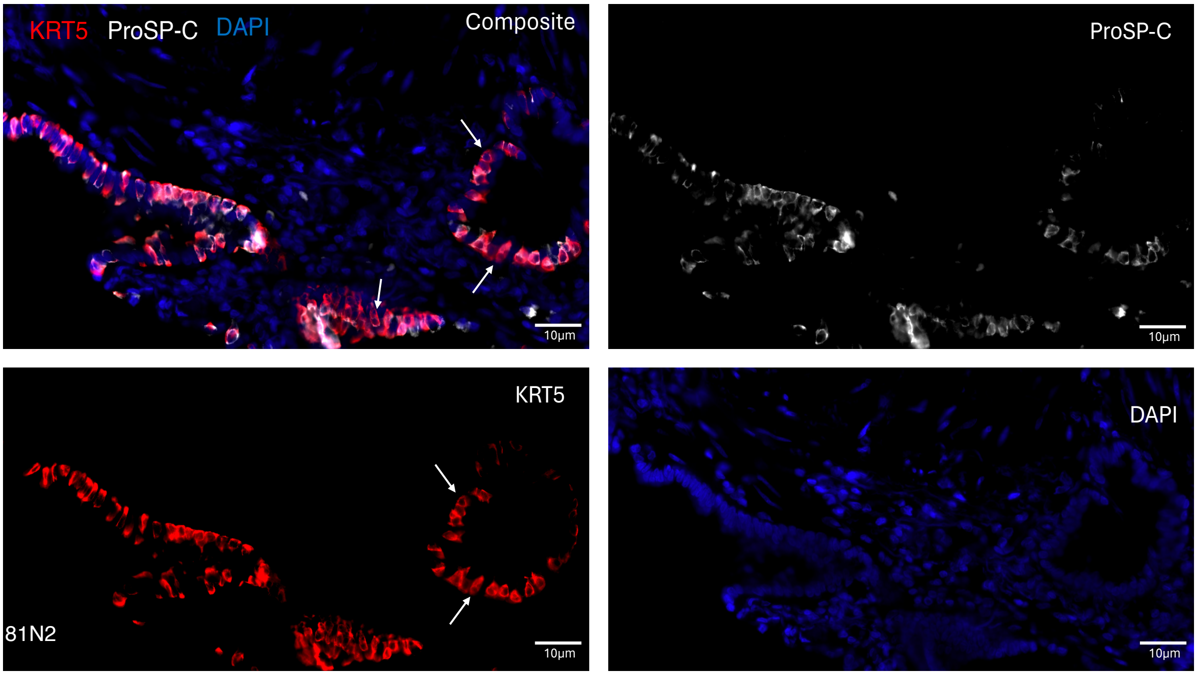

White arrows – Single positive staining - KRT5`+ (Basal cells). All other cells double positive for KRT5 and ProSp-C (ABI_a and ABI_b)

**Suppl Figure 3**

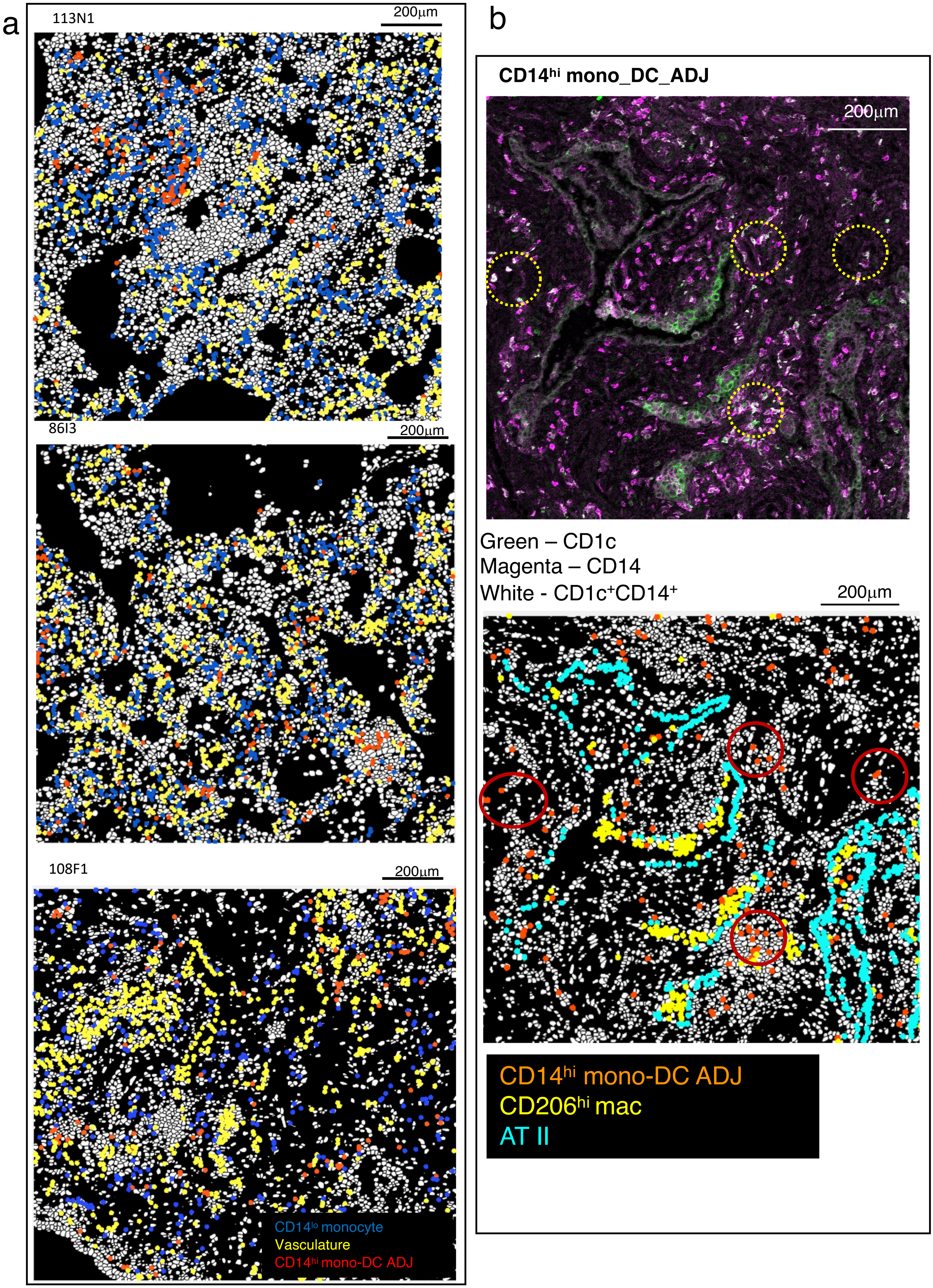

**Suppl Figure 4**

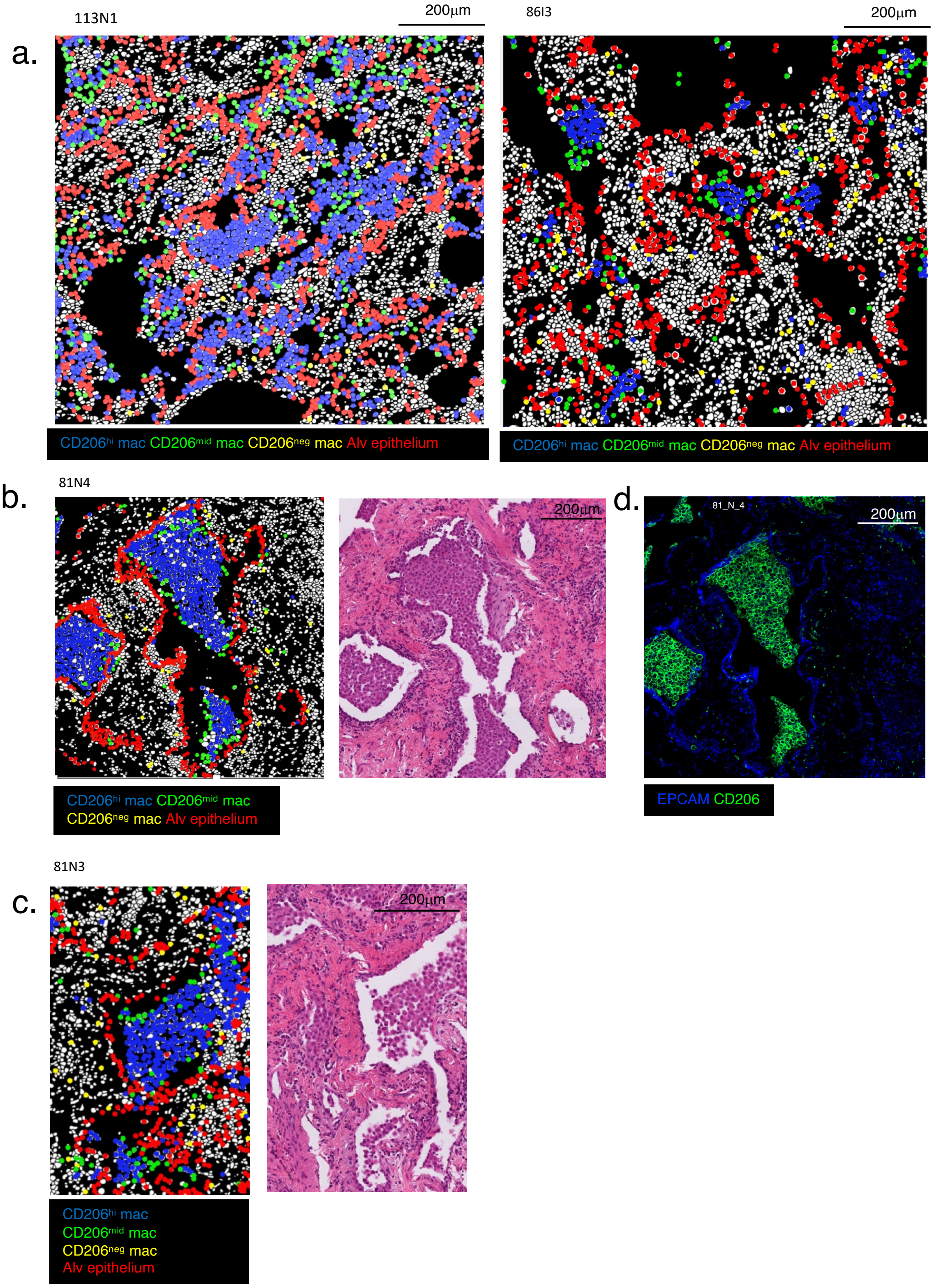

**Suppl Figure 5**

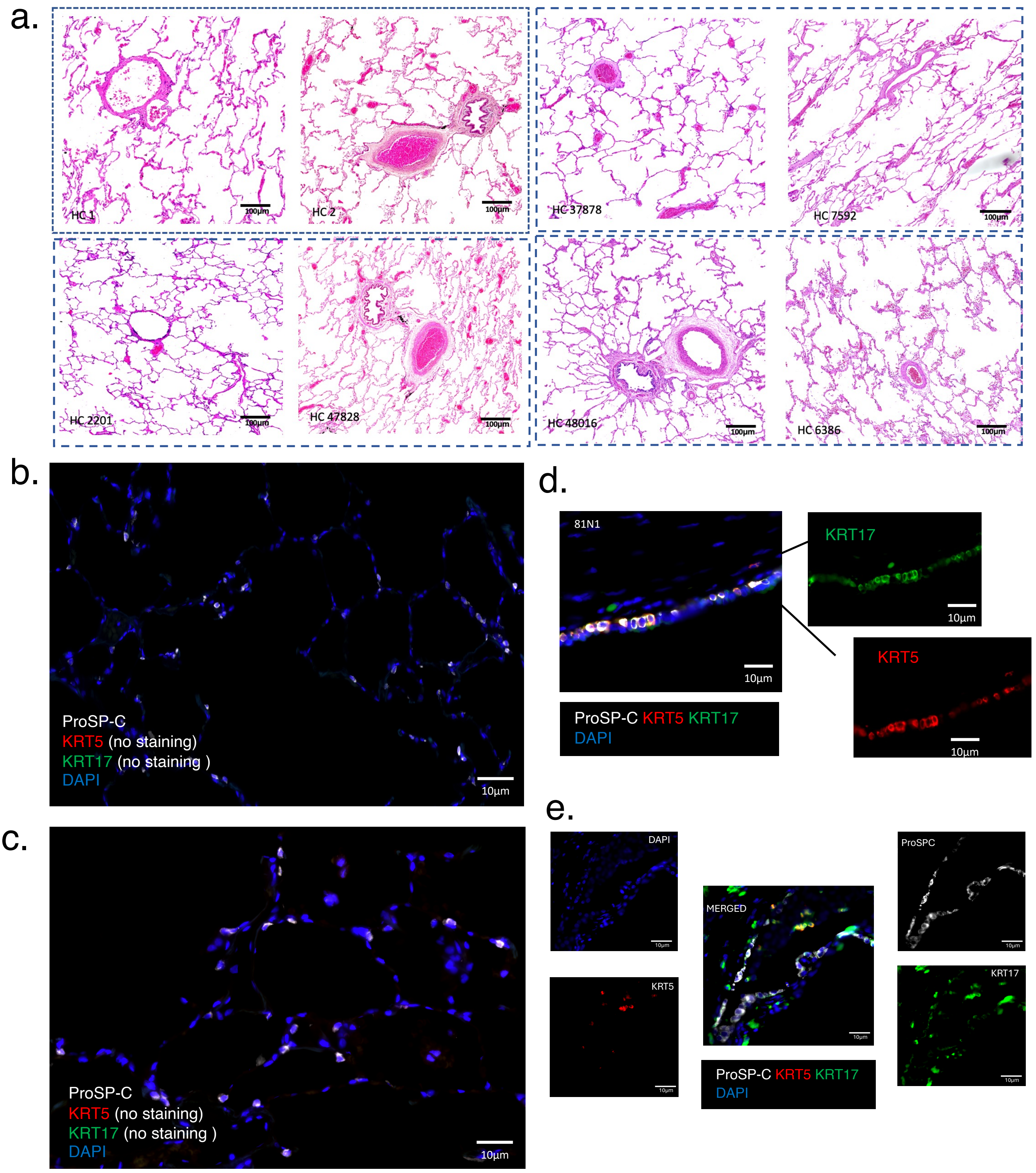

**Suppl Figure 6**

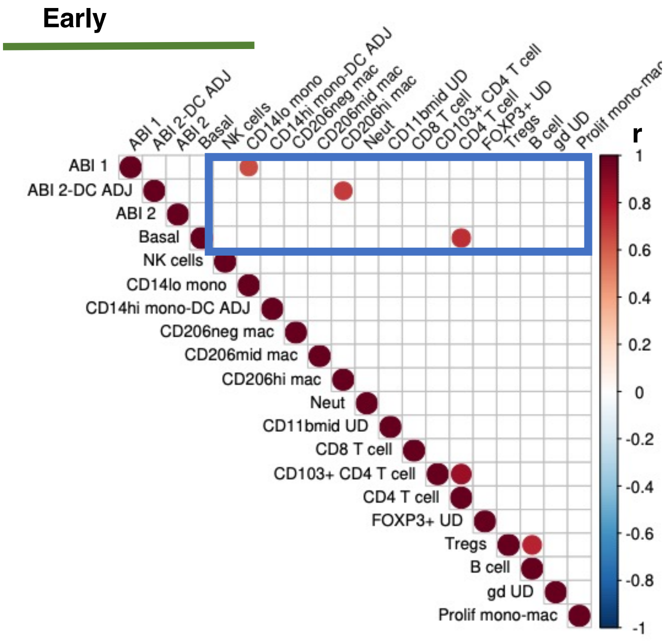

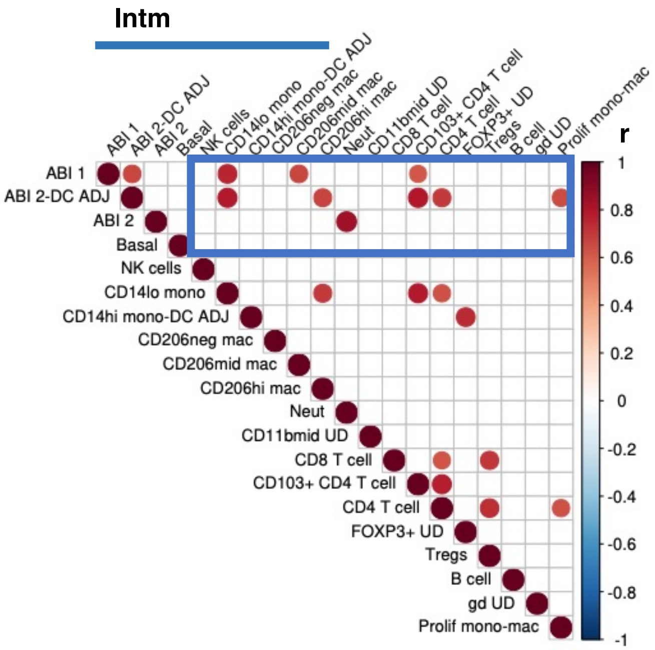

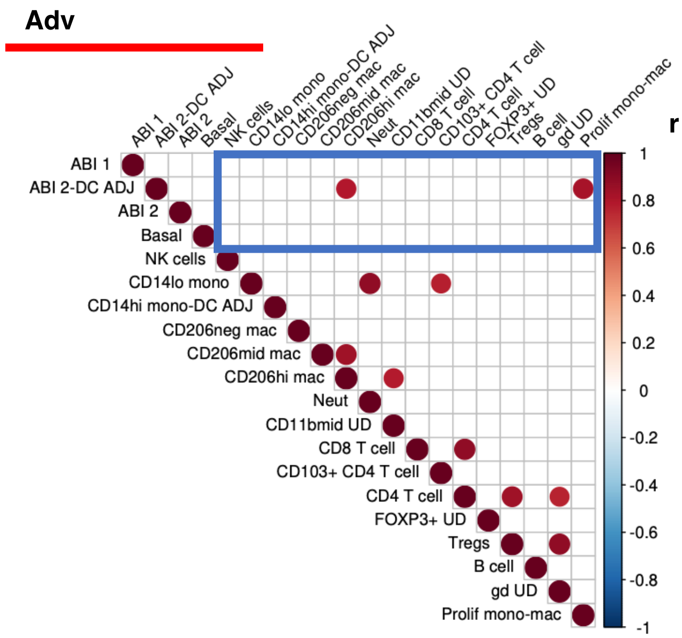

**Suppl Figure 7**

a.

### Cross PCF $g_{C_1C_2}(r)$ for cells of types $C_1$ and $C_2$

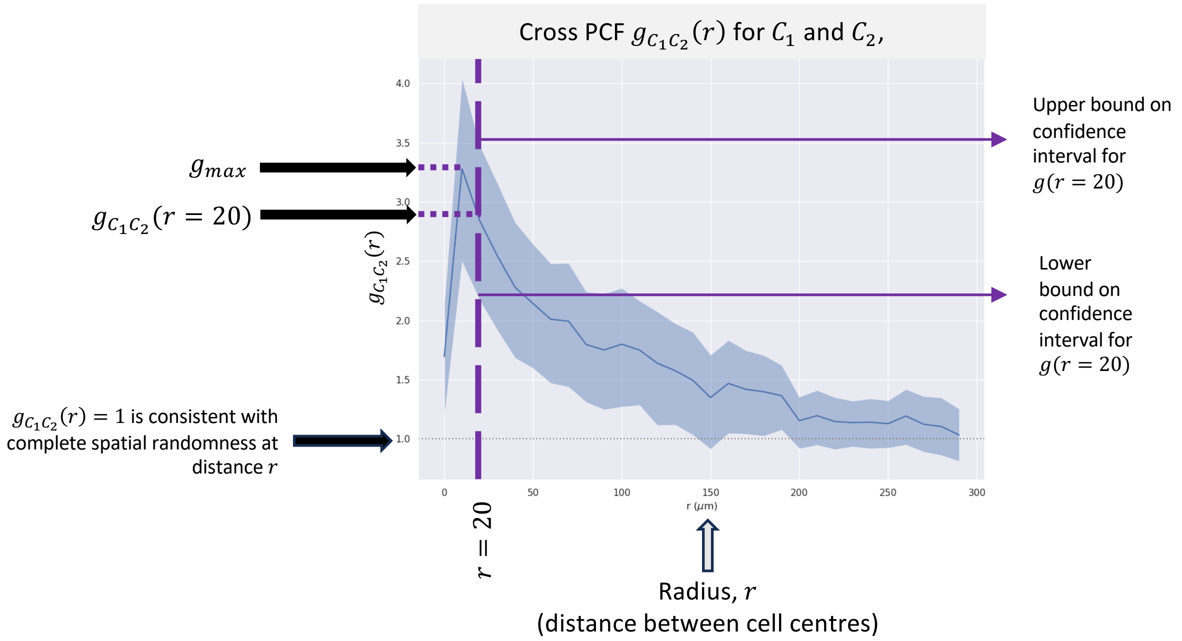

$$g_{C_1C_2}(r) = \frac{1}{N_{C_1}} \sum_{i=1}^{N} \mathbb{I}(C_1, c_i) \left( \sum_{j=1}^{N} \mathbb{I}(C_2, c_j) \frac{I_{[0,dr)}(|\boldsymbol{x}_i - \boldsymbol{x}_j| - \boldsymbol{r})}{A_r(\boldsymbol{x}_i)} \Big/ \frac{N_{C_2}}{A} \right)$$

where:

- $N_{C_i} = \sum_{j=1}^{N} \mathbb{I}(C_i, c_j)$
- $\mathbb{I}(C, c_i) = 1$ if $c_i = C$, or 0 otherwise
- $I_{[a,b)}(r) = 1$ if $r \in [a, b)$, or 0 otherwise (for $a < b$ and $a, b \in \mathbb{R}$)

Suppl Figure 8A

b.

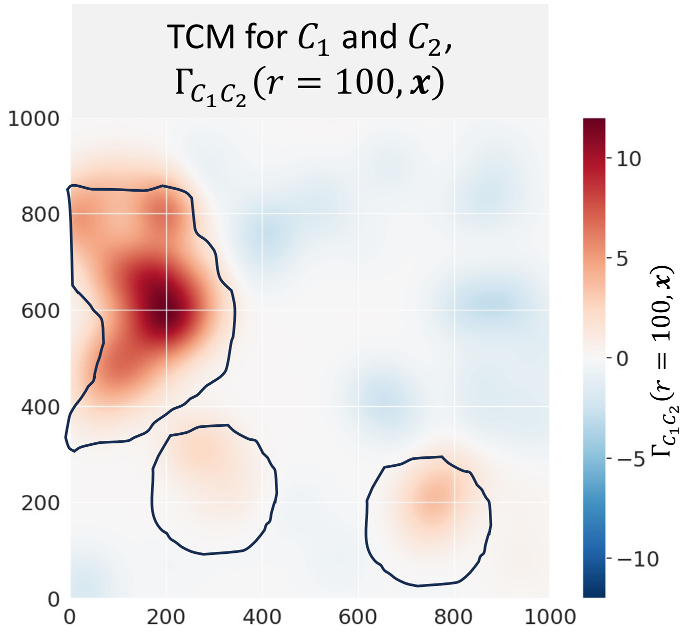

Highlighted regions depict co–location of $C_1$ and $C_2$ cells

$$\Gamma_{C_1C_2}(r, \boldsymbol{x}) = \sum_{i=1}^{N_{C_1}} \frac{\mu_{C_1C_2}}{2\pi\sigma^2} e^{-\frac{1}{2}\left(\frac{|\boldsymbol{x}-\boldsymbol{x}_i|}{\sigma}\right)^2}$$

where:

- $\boldsymbol{x}_i$ is the coordinate of cell $i$
- $\mu_{C_1C_2}(r, \boldsymbol{x}_i)$ is a normalised representation of the contribution of cell $i$ to the cross-PCF at radii in the range from 0 to $r$
- $\sigma$ is a parameter relating to the smoothness of the TCM

**Suppl Figure 8B**

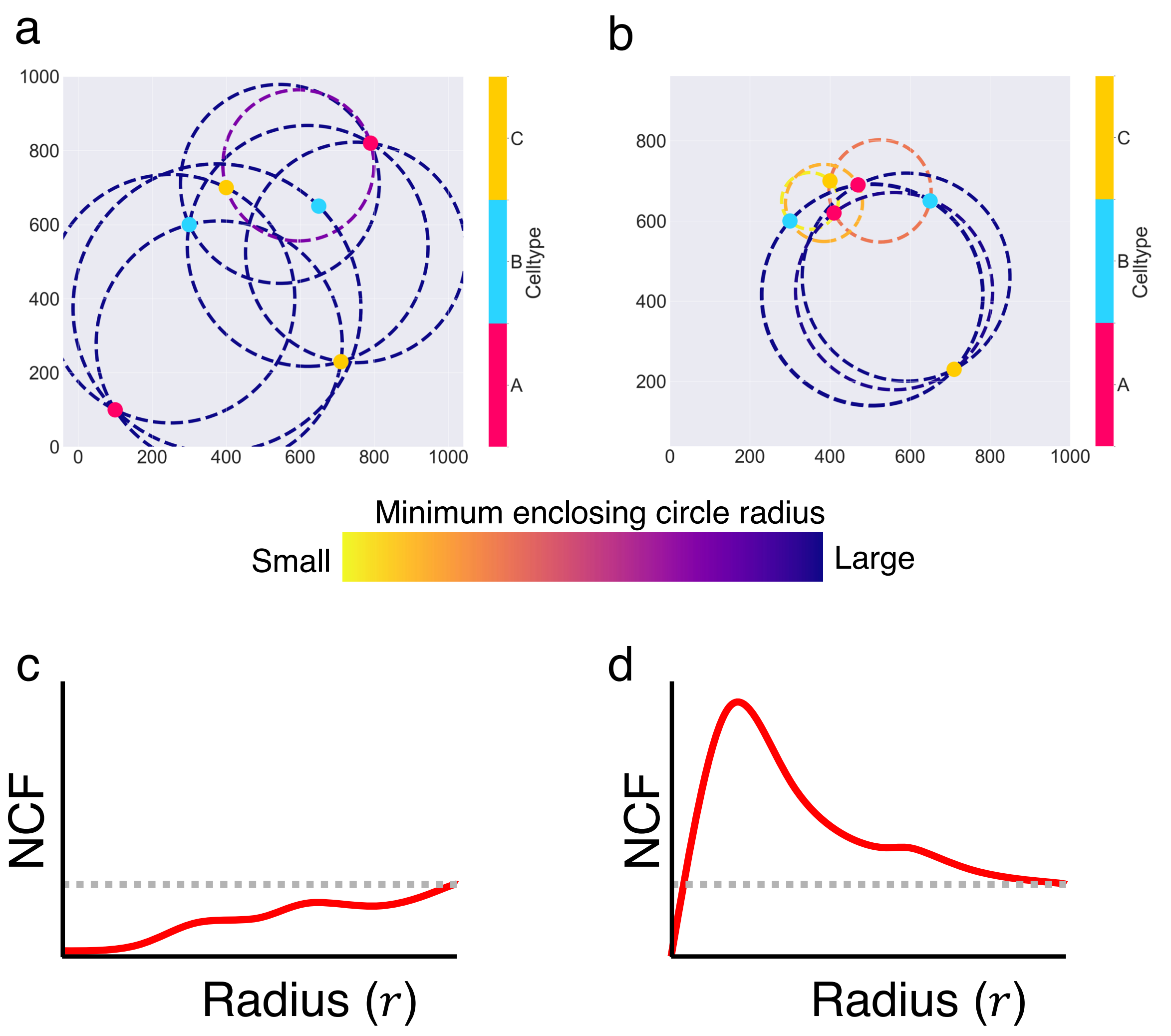

**Suppl Figure 9**

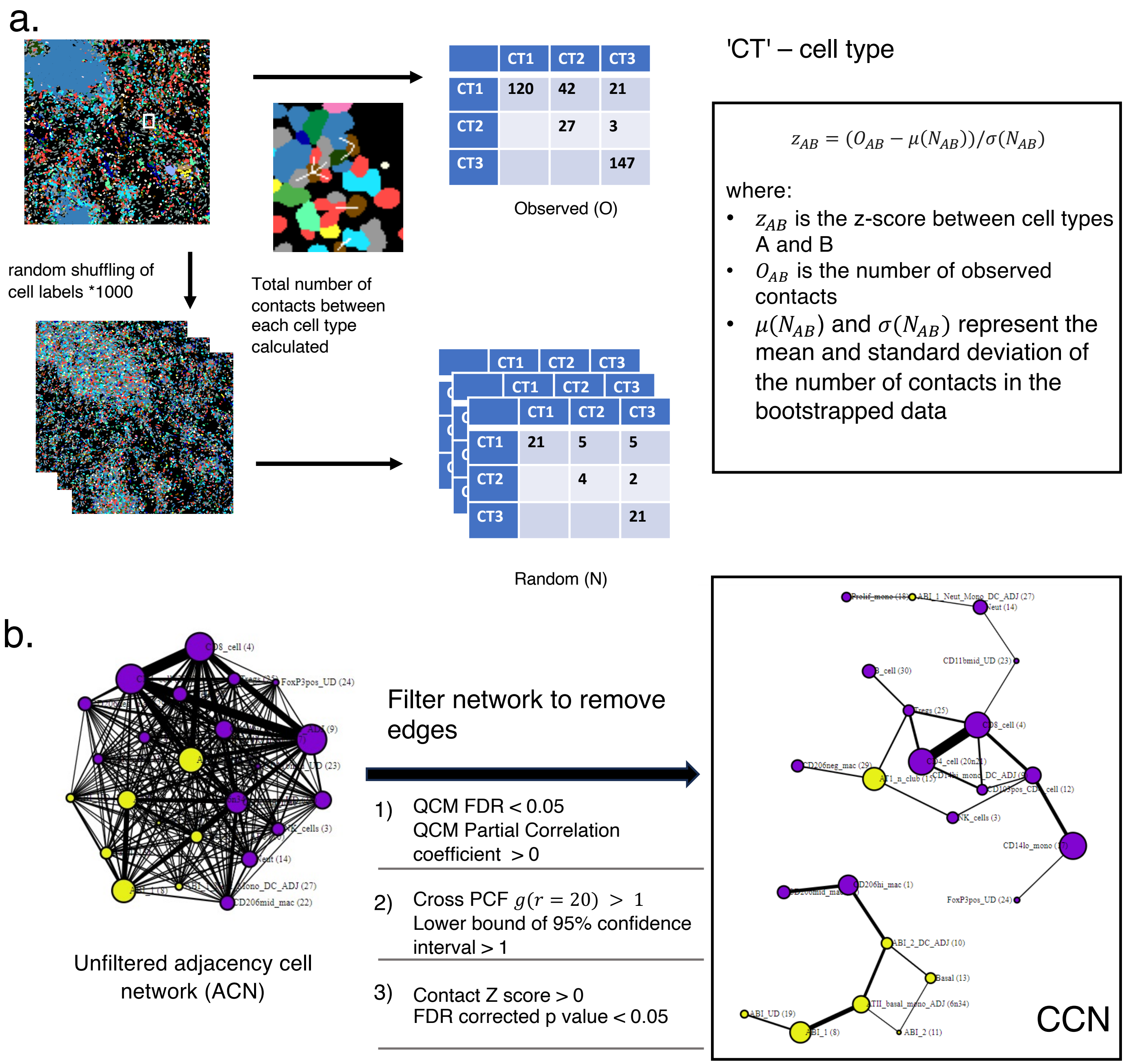

**Suppl Figure 10**

| Patient ID | Diagnosis | Sex | Age | Lung function tests | | |
|---|---|---|---|---|---|---|
| | | | | **FEV1(% of predicted)** | **FVC (% of predicted)** | **TLCO (% of predicted)** |
| RB81 | IPF | Male | 64 | 2.29 (64%) | 3.02 (66%) | 2.87(28%) |
| RB82 | IPF | Female | 59 | 2.10 (82%) | 2.44(81%) | 1.70(21%) |
| RB86 | IPF | Male | 55 | 2.83(91%) | 3.42(89%) | 2.12(24%) |
| RB98 | IPF | Male | 61 | 2.10(64%) | 2.32(56%) | 1.76(19%) |
| RB104 | IPF | Female | 50 | 1.33(46%) | 1.51(45%) | 2.21(25%) |
| RB108 | IPF | Male | 63 | 2.61(59%) | 3.30(54%) | 1.46(19%) |
| RB113 | IPF | Male | 50 | 2.10(56%) | 2.55(55%) | 2.45(23%) |
| RB051 | HC | Male | 28 | N/A | N/A | N/A |
| RB052 | HC | Male | 42 | N/A | N/A | N/A |

**Supplementary Table 1**. Demographic of patients at the point of sample harvesting.
N/A – Not available

| Disease Stage and ROI identifier | cell_number | sample_id | Overall histopathology picture * | Fibroblastic foci (FF) (0=none; 1=present) | Number of FF | Bronchiolisation (0=none; 1=present) | |
|---|---|---|---|---|---|---|---|
| **Advanced** | | | | | | | |
| 25 | 5695 | IPF_SAMPLE_081F_ROI_1 | C | **0** | 0 | 0 | **n=14 ROI** |
| 33 | 5493 | IPF_SAMPLE_081F_ROI_2 | A/B | **0** | 0 | 0 | **10 C, 2 B/C, 1 A/B, 1 B** |
| 12 | 4915 | IPF_SAMPLE_086F_ROI_1 | C | **0** | 0 | 0 | **71%C, 14% B/C** |
| 45 | 5011 | IPF_SAMPLE_086F_ROI_2 | C | **0** | 0 | 0 | **14% of ROIs have FF** |
| 44 | 10198 | IPF_SAMPLE_098F_ROI_1 | B/C | **1** | 1 | 0 | **total cells = 89826** |
| 26 | 11193 | IPF_SAMPLE_104F_ROI_2 | C | **0** | 0 | 0 | |
| 18 | 4477 | IPF_SAMPLE_108F_ROI_1 | C | **0** | 0 | 1 | |
| 48 | 6344 | IPF_SAMPLE_113F_ROI_1 | C | **1** | 2 | 0 | |
| 55 | 6663 | IPF_SAMPLE_113F_ROI_2 | C | **0** | 0 | 0 | |
| 11 | 5252 | IPF_SAMPLE_113F_ROI_3 | C | **0** | 0 | 0 | |
| 0 | 5390 | IPF_SAMPLE_113F_ROI_4 | C | **0** | 0 | 0 | |
| 59 | 7416 | IPF_SAMPLE_113F_ROI_5 | C | **0** | 0 | 0 | |
| 42 | 5080 | IPF_SAMPLE_82F_ROI_2 | B | **0** | 0 | 0 | |
| 27 | 6699 | IPF_SAMPLE_82F_ROI_3 | B/C | **0** | 0 | 1 | |
| **Intm** | | | | | | | |
| 35 | 6179 | IPF_SAMPLE_081I_ROI_1 | B | **1** | 2 | 0 | **n=20 ROI** |
| 3 | 4989 | IPF_SAMPLE_081I_ROI_2 | B | **1** | 1 | 0 | **2 C, 4 B/C, 9 B, 2 A/B, 3A** |
| 46 | 6388 | IPF_SAMPLE_081I_ROI_3 | B | **1** | 2 | 0 | **45%B, 20% B/C** |
| 56 | 4977 | IPF_SAMPLE_081I_ROI_4 | B | **0** | 0 | 0 | **55% has FF** |
| 57 | 4720 | IPF_SAMPLE_086I_ROI_1 | A | **1** | 3 | 0 | **total cells = 140278** |
| 20 | 4724 | IPF_SAMPLE_086I_ROI_2 | A | **1** | 3 | 0 | |
| 50 | 4991 | IPF_SAMPLE_086I_ROI_3 | A | **0** | 0 | 0 | |
| 43 | 9858 | IPF_SAMPLE_098I_ROI_1 | B | **1** | 3 | 0 | |
| 60 | 9783 | IPF_SAMPLE_098I_ROI_2 | B | **1** | 2 | 0 | |
| 14 | 10026 | IPF_SAMPLE_098I_ROI_3 | A/B | **0** | 0 | 0 | |

| | | | | | | | |
|---|---|---|---|---|---|---|---|
| 34 | 9775 | IPF_SAMPLE_098I_ROI_4 | B/C | **1** | 1 | 0 | |
| 32 | 9841 | IPF_SAMPLE_098I_ROI_5 | B/C | **1** | 3 | 0 | |
| 39 | 9852 | IPF_SAMPLE_104I_ROI_1 | B/C | **0** | 0 | 0 | |
| 36 | 7734 | IPF_SAMPLE_104I_ROI_2 | B | **1** | 1 | 1 | |
| 16 | 6127 | IPF_SAMPLE_108I_ROI_1 | C | **0** | 0 | 0 | |
| 23 | 6410 | IPF_SAMPLE_108I_ROI_2 | B/C | **0** | 0 | 0 | |
| 61 | 4186 | IPF_SAMPLE_113I_ROI_1 | A/B | **1** | 1 | 0 | |
| 24 | 7717 | IPF_SAMPLE_113I_ROI_2 | B | **0** | 0 | 0 | |
| 29 | 6157 | IPF_SAMPLE_113I_ROI_3 | B | **0** | 0 | 0 | |
| 10 | 5844 | IPF_SAMPLE_113I_ROI_5 | B/C | **0** | 0 | 0 | |
| **Early** | | | | | | | |
| 22 | 4289 | IPF_SAMPLE_081N_ROI_1 | C | **0** | 0 | 0 | **n=19 ROI** |
| 53 | 5104 | IPF_SAMPLE_081N_ROI_2 | C | **0** | 0 | 0 | **5 C, 5 B/C, 1B, 3 A/B, 5A** |
| 38 | 6305 | IPF_SAMPLE_081N_ROI_3 | B/C | **0** | 0 | 0 | **26% A, 26%B/C, 26% C** |
| 9 | 5715 | IPF_SAMPLE_081N_ROI_4 | B/C | **1** | 1 | 0 | **47% has FF** |
| 6 | 7602 | IPF_SAMPLE_098N_ROI_1 | C | **0** | 0 | 0 | **total cells = 112406** |
| 2 | 9575 | IPF_SAMPLE_098N_ROI_2 | C | **0** | 0 | 0 | |
| 52 | 7256 | IPF_SAMPLE_098N_ROI_3 | B | **1** | 2 | 0 | |
| 8 | 7033 | IPF_SAMPLE_108N_ROI_1 | M/L | **1** | 1 | 0 | |
| 47 | 5603 | IPF_SAMPLE_108N_ROI_2 | C | **0** | 0 | 0 | |
| 30 | 6599 | IPF_SAMPLE_113N_ROI_1 | A | **0** | 0 | 0 | |
| 40 | 7194 | IPF_SAMPLE_113N_ROI_2 | A | **0** | 0 | 0 | |
| 17 | 6307 | IPF_SAMPLE_113N_ROI_3 | A/B | **1** | 3 | 0 | |
| 7 | 3938 | IPF_SAMPLE_82N_ROI_1 | A/B | **1** | 2 | 0 | |
| 41 | 4410 | IPF_SAMPLE_82N_ROI_2 | B/C | **1** | 2 | 0 | |
| 49 | 3726 | IPF_SAMPLE_82N_ROI_3 | B/C | **0** | 0 | 0 | |
| 13 | 4422 | IPF_SAMPLE_82N_ROI_4 | A/B | **1** | 2 | 0 | |
| 19 | 5315 | IPF_SAMPLE_86N_ROI_1 | A | **0** | 0 | 0 | |
| 51 | 7120 | IPF_SAMPLE_86N_ROI_2 | A | **1** | 3 | 0 | |
| 54 | 4893 | IPF_SAMPLE_86N_ROI_4 | A | **1** | 1 | 0 | |

**Supplementary Table 2. Histopathology analysis of lung sections.**

***Overall histopathology picture**

a. Collagen less prominent, more fibroblastic foci (FF), more inflammatory infiltrate, less expanded interstitium

b. More collagen, less FF, inflammatory infiltrate, more expanded interstitium

c. Most collagen, more prominent smooth muscle hyperplasia, v little airspace left, little or no FF, occasional evidence of bronchiolisation

| Marker | Metal |
|---|---|
| CD45 | 141Pr |
| CD68 | 142Nd |
| CD8a | 143Nd |
| Ki67 | 144Nd |
| alpha-Smooth Muscle Actin | 145Nd |
| FoxP3 | 147Sm |
| CD11c | 149Sm |
| CD103 | 152Sm |
| CD56 | 153Eu |
| Helios | 154Sm |
| CD69 | 155Gd |
| Epcam | 156Gd |
| CD20 | 158Gd |
| CD206 | 159Tb |
| CD127 | 160Gd |
| KRT17 | 161Dy |
| CD1c | 162Dy |
| CD15 | 163Dy |
| KRT14 | 164Dy |
| TCRγ | 165Ho |
| Amphiregulin | 166Er |
| CD11B | 167Er |
| BDCA2 | 168Er |
| KRT5 | 169Tm |
| CD3 | 170Er |
| proSP -C | 171Yb |
| CD31 | 172Yb |
| CD161 | 173Yb |
| CD4 | 174Yb |
| HLA-DR | 175Lu |
| CD14 | 176Yb |
| DNA - 1 | 191Ir |
| DNA - 3 | 193Ir |

**Supplementary table 3 – 33 plex panel with their metal tags for Imaging Mass Cytometry.**

A.

| % of all cells | Cluster | Name | Description and expanded notes |
|---|---|---|---|
| 1.60% | 3 | **NK cells** | $CD56^{+}$ population. |
| 10.30% | 17 | **$CD14^{lo}$ mono** | All cells are $CD14^{lo}$ and HLADR $^{mid}$ and CD68 $^{neg.}$ Likely patrolling monocytes found on or near endothelium of blood vessels(*1*). Spatial analysis in our work shows co-location with vasculature. |
| 3.10% | 9 | **$CD14^{hi}$ mono-DC ADJ** | $CD14^{hi}$ – the only $CD14^{hi}$ cluster - classical monocytes ($CD14^{hi}CD16^{neg}$ monocytes)(*2*). Likely found with DC adjacent to it, due to $CD11c^{hi}$, $CD1c^{hi}$ and $HLADR^{hi}$ expression. Can also be transitional macrophages (transitioning between monocytes to macrophage in differentiation pathway)(*2*). See also Suppl Figure 4B for staining. |
| 1.30% | 29 | **$CD206^{neg}$ mac** | $CD68^{lo}$ $CD206^{neg}CD14$ $^{neg}$ HLADR $^{neg}$; likely interstitial macrophage due to location of cells. See Suppl Fig 5A-C for location in cell centroid maps. |
| 1.60% | 22 | **$CD206^{mid}$ mac** | $CD68^{mid}CD206^{mid}CD14^{neg}HLADR^{mid}$; alveolar macrophages. Always found with $CD206^{hi}$ macrophages in alveolar lumen; note CD68 expression lower suggesting earlier in differentiation trajectory from monocyte to macrophages but they are $CD14^{neg}$. Spatial co-location with $CD206^{hi}$ mac supports this possibility of less mature alveolar macrophage (Fig 5D-F). See Suppl Fig 5A-C for location in cell centroid maps. |
| 5.90% | 1 | **$CD206^{hi}$ mac** | $CD68^{hi}$ $CD206^{hi}$ $CD14^{mid}$ $HLADR^{hi}$. Alveolar macrophages. Most mature macrophage with high HLA DR and CD68. See Suppl Fig 5A-C for location in cell centroid maps. |
| 3.70% | 14 | **Neut** | Neutrophils. Only CD15 expressing cells found |
| 0.30% | 23 | **$CD11b^{mid}$ UD** | Clear CD11b expression but no other defining features. Identity unknown. |
| 6.90% | 4 | **CD8 T cell** | CD69 $^{neg}$ CD11b $^{neg}$ CD103 $^{neg}$ CD8 T cells |
| 1.10% | 12 | **$CD103^{+}$ CD4 T cell** | CD68 $^{neg}$ $CD103^{+}$ CD4 T cells. Likely resident CD4 T cells; usually found in alveolar lining. |
| 7.10% | 20 | **CD4 T cell** | CD69 $^{neg}$ CD4 T cells |
| 2.20% | 21 | **CD4 T cell** | CD69 neg CD4 T cells |
| 0.30% | 24 | **$FOXP3^{+}$ UD** | FOXP3 $^{mid-hi}$ CD3 $^{neg}$ CD4 $l^{o}$ cells $HELIOS^{mid}$ $AREG^{lo.}$ Identity unclear. |
| 1.10% | 25 | **Tregs** | FOXP3 $^{lo-mid}$ CD3 $^{mid-hi}$ CD4 $^{mid\ -hi}$ HELIOS $^{mid}$ AREG $^{lo}$ |
| 4.50% | 30 | **B cell** | CD20 $^{mid-hi}$ |
| 3% | 26 | **γδ UD** | CD45 $^{neg}$ and EPCAM $^{neg}$, could be non-specific staining |
| 1.20% | 18 | **Prolif mono-mac** | Proliferating monocyte-macrophage. Almost identical to cluster 17 ($CD14^{lo}$ mono) - all cells are $CD14^{lo}$ , HLADR $^{mid}$ and Ki67 $^{mid\ -hi}$. Only cluster to show this high level of Ki67 expression |
| 5.50% | 15 | **15 UD** | $CD45^{lo}$, $EpCAM^{lo}$, $ProSPC^{neg}$, $KRT5^{neg}$, $KRT17/7^{neg}$ - spatial check on cell centroid map and IMC images suggest these are immune cells found scattered in interstitium. Some cells found close to ABIs. Likely immune cells with some cells adjacent to ABIs. Included in immune cell group. Unlikely to contain ATI as 15 UD cluster is $KRT17/7^{neg}$ while Haberman's dataset (Suppl Fig 3B clearly shows ATI expressing high levels of KRT 7) |

| | | | |
|---|---|---|---|
| 10.10% | 5 | **Vasculature** | CD31$^{mid}$, $\alpha$SMA $^{neg-lo-mid}$. A mix of arterioles, venules and lymphatics |
| 0.20% | 28 | **Endothelium** | CD31$^{lo}$, mainly capillaries in alveolar bed |
| 5.40% | 2 | **SM** | Smooth muscle. $\alpha$SMA$^{hi}$ CD45$^{neg}$ and EPCAM $^{neg}$ - could be fibroblasts, smooth muscle cells as part of SM hyperplasia in IPF or smooth muscle around arteries or bronchioles |
| 4.90% | 6 | **ATII** | AT II cells (high proSP-C expressing cells); mainly KRT5$^{neg}$, KRT17/7 positive on IMC (likely due to KRT7 positivity). Immunofluorescence staining shows negative KRT17 staining (Suppl Fig 3E) Final Cluster 6 included both cluster 6 and 34 |
| 5.30% | 8 | **ABI_a** | Early ABI in trajectory between KRT5$^{-}$KRT17/7$^{+}$ ABI and basal cells |
| 1.90% | 10 | **ABI b-DC ADJ** | Mid ABI in trajectory between KRT5$^{-}$KRT17/7$^{+}$ ABI and basal cells, adjacent to DCs which can be in lumen or interstitium; note it is the only ABI with low CD45 expression in keeping with presence of immune cell (DC) (Suppl Fig 2A; Fig 2F). Does not discriminate between DC and CD11c-expressing macrophages but called DC due to higher expression of CD1c and lower expression of CD68 and CD206 (see Suppl Table 4B below). |
| 0.40% | 11 | **ABI_b** | Mid ABI in trajectory between KRT5$^{-}$KRT17/7$^{+}$ ABI and basal cells. Note very small numbers of cells. |
| 1.70% | 13 | **Basal** | KRT5$^{hi}$ proSP-C$^{neg}$ cells. Likely end point of ABI differentiation trajectory. Staining shown in Suppl Fig 3F |
| 1.30% | 19 | **19 UD** | EpCAM$^{neg}$, CD45$^{neg}$, KRT5$^{lo}$, KRT17/7 $^{neg/lo}$ - identity unclear |
| 0.40% | 27 | **27 UD** | EpCAM $^{neg-lo}$ ,ProSP-C $^{lo}$, KRT5 $^{neg-lo}$, KRT17/7$^{mid}$, CD14$^{lo}$, BDCA2$^{mid}$, CD11b$^{mid}$,CD15 $^{mid}$, CD14$^{lo}$, CD1c $^{lo-mid}$. Likely mix of immune cells (neutrophils, monocytes, DC) with an ABI , ATI or AT II. Very small numbers of cells. Spatial co-location shows spatial association with Neutrophils in all disease stages (Fig 5D-F). Included in epithelial cell group |
| 0.04% | 34 | **ATII** | AT II cells (high ProSP-C expressing cells) – merge with Cluster 6 and overall called Cluster 6. |

1. C. Auffray et al., Monitoring of Blood Vessels and Tissues by a Population of Monocytes with Patrolling Behavior. Science 317, 666-670 (2007).
2. E. Fraser et al., Multi-Modal Characterization of Monocytes in Idiopathic Pulmonary Fibrosis Reveals a Primed Type I Interferon Immune Phenotype. Front Immunol 12, 623430 (2021).

B.

| Myeloid cell clusters and selected epithelial cells and their relative expression of relevant markers | | | | | | | | |
|---|---|---|---|---|---|---|---|---|
| **Name** | **Cluster number** | **CD11c** | **CD14** | **CD1c** | **HLADR** | **CD68** | **CD206** | **BDCA2** |
| **CD206$^{hi}$ mac** | 1 | + | ++ | +++ | +++ | +++ | +++ | + |
| **CD14$^{hi}$ mono-DC ADJ** | 9 | ++ | +++ | +++ | +++ | +/- | - | + |
| **CD14$^{lo}$ mono** | 17 | - | ++ | + | ++ | - | - | - |
| **Prolif mono-mac** | 18 | - | ++ | + | ++ | - | - | + |
| **CD206$^{mid}$ mac** | 22 | + | - | + | ++ | ++ | ++ | - |
| **CD206$^{neg}$ mac** | 29 | - | - | - | - | +/- | - | - |
| **ABI b-DC ADJ** | 10 | + | ++ | +++ | +++ | +/- | +/- | + |
| **Basal** | 13 | - | ++ | +++ | +/- | - | - | + |
| **27 UD** | 27 | - | ++ | + | +/++ | - | - | + |
| **ATII** | 6 | - | ++ | + | +++ | - | - | + |

**Supplementary Table 4. Annotation of clusters, and expanded notes on annotation.**
